# Paving the way for automated transscleral cyclophotocoagulation: predicting ciliary body arc length from biometric data using a two-sphere eye model

**DOI:** 10.64898/2026.03.29.26349517

**Authors:** Áron Szabó, Tamás Árpádffy-Lovas, Edit Tóth-Molnár

## Abstract

**Purpose:** To improve determination of the treatment area for the personalization of subliminal transscleral cyclophotocoagulation (SL-TSCPC) procedures in glaucoma treatment, we designed a biometry based model of the human eye to find the estimated cilary body (CB) arc length (ECBAL) and the calculated CB distance (CCBD).

**Methods:** We developed a rotationally symmetric modified two-sphere eye model based on axial length (AL), mean keratometry (mean K), anterior chamber depth (ACD), lens thickness (LT), and white-to-white (WTW). ECBAL and CCBD were calculated for each eye. Fluence was calculated with standardized parameters.

**Results:** Publicly accessible biometric measurements for 24,001 eyes were pooled for analysis. The mean ECBAL was 23.99±1.8 mm. The correlations of ECBAL with AL and ACD were 0.723 and 0.754 respectively (p < 0.01). The number of eyes with an ECBAL 21.7–22.0 mm was 131 of 24,001 (0.55%). The mean CCBD was 4.21±0.8 mm. The number of eyes with a CCBD of 3.8 mm was 1,445 of 24,001 (6.02%). Mean fluence was 120.33±9.0 J/cm2. A mean difference of −8.18 ± 6.9%, ranging from −22.66% to +29.07% in fluence was observed with treating only the recommended 22 mm versus the ECBAL.

**Conclusions:** This study demonstrated that use of 22.0 mm as the standard treatment arc length may under or overdose laser treatment in many eyes. Precise estimation or exact localization of the CB treatment area is required to accurately calculate fluence.

**Translational Relevance:** The model proves that CB arc length is a variable while current guidelines consider it a constant

## Introduction

Reducing intraocular pressure (IOP) is currently the only way to slow the progression of glaucoma. This can be achieved either by decreasing aqueous production or increasing outflow.^1^ Various pharmacological agents, surgical techniques and devices, and laser procedures have been developed to reduce IOP while maintaining an acceptable cost-benefit profile. Recent advances in subliminal transscleral cyclophotocoagulation (SL-TSCPC) have made this a safe and efficient surgical pathway.^2^ SL-TSCPC delivers repetitive pulses of 810 nm light toward the ciliary body (CB) using a pen-like device held by the surgeon. This infrared light raises the temperature of the target tissue, which leads to a decrease in aqueous production by directly inducing apoptosis and an increase in outflow by inducing shrinkage of the ciliary body. In contrast to continuous treatment, repetitive laser pulsation has pauses between pulses. This limits the extent of local tissue damage by allowing the tissue to cool between pulses and has been shown to reduce adverse events.^3^ Numerous variables affect the exact amount of energy that reaches the target tissue. The most widely utilized SL-TSCPC systems, such as the IRIDEX MicroPulse® and Quantel Vitra/Supra 810 platforms, allow the operating surgeon to adjust both the power delivered with each application and the duty cycle. The duty cycle refers to the amount of time the laser spends in the ON position and delivers energy, with the rest of the time spent in the OFF position. The total exposure time refers to the overall duration of laser treatment. The probes can deliver the laser treatment with varying surface areas, which currently ranges between 600 and 700 µm.

Because the laser pulses are targeted toward the tissue by hand, the surgeon’s movements can influence treatment efficacy. Even when the total exposure time and power are constant, variation in the time spent over a particular area can result in marked changes in the amount of energy that reaches the tissue. This variable is referred to as the dwell time. Dwell time is associated with the velocity of the probe moving on the surface of the conjunctiva, which is the sweep velocity. Less time spent over an area will result in a lower amount of energy delivered to that area if the increase in sweep velocity is not corrected for by increasing the power. The concept of fluence was introduced to correct for probe movement. Fluence is expressed in J/cm^2^ and represents the energy delivered to each segment of the target tissue. Calculation of fluence requires knowing the power, duty cycle, total exposure time, probe surface area, and sweep time or velocity. As shown by prior studies, fluence may predict the expected IOP-lowering effect of SL-TSCPC treatment better than the total delivered energy.^4^

The 2022–2023 consensus guidelines on SL-TSCPC ^5,6^ provide reference values for power (2500 mW), duty cycle (31.3%), sweep time (20 seconds), and the number of sweeps (4) per hemisphere. These values correspond to a total energy of 125.2 J provided that both hemispheres are treated and the fluence is 131 J/cm^2^. Because sweep velocity can vary substantially with subtle changes in the size and position of the CB, exact calculation of fluence requires precise measurements of the dimensions of the target area (arc length).

In the appendix of the original fluence calculation formula paper^4^, the probe was positioned 3.8 mm posterior to the limbus and the arc length of treatment was 21.7 mm. In subsequent works, the latter value is rounded to 22.0 mm and used as a constant. However, the dimensions of the treated eye were not discussed.

Automation of laser procedures is an ongoing development in ocular surgery, as reflected by advances in corneal refractive procedures^7^, direct selective laser trabeculoplasty (dSLT)^8^, and femtosecond laser image-guided trabeculotomy (FliGHT).^9^ Although targeting of the CB in SL-TSCPC is currently done manually, incorporating a variable-length treatment area could enable more precise energy delivery, improve standardization and personalization, and facilitate future automation.

This study aimed to design an algorithm based on biometric data for the estimated CB arc length (ECBAL) and the distance of this plane from the white-to-white (WTW), which we term the calculated CB distance (CCBD). Using our theoretical model, we aimed to improve determination of the treatment area, approximate the size of the reference eye, and evaluate the effect of individual eye anatomy on fluence using a large publicly available dataset.

## Methods

### Theoretical model construction

We developed a rotationally symmetric modified two-sphere model (Figure 1) based on the following biometric parameters of the eye: axial length (AL), mean keratometry (mean K), anterior chamber depth (ACD), lens thickness (LT), and WTW.

**Figure 1.**
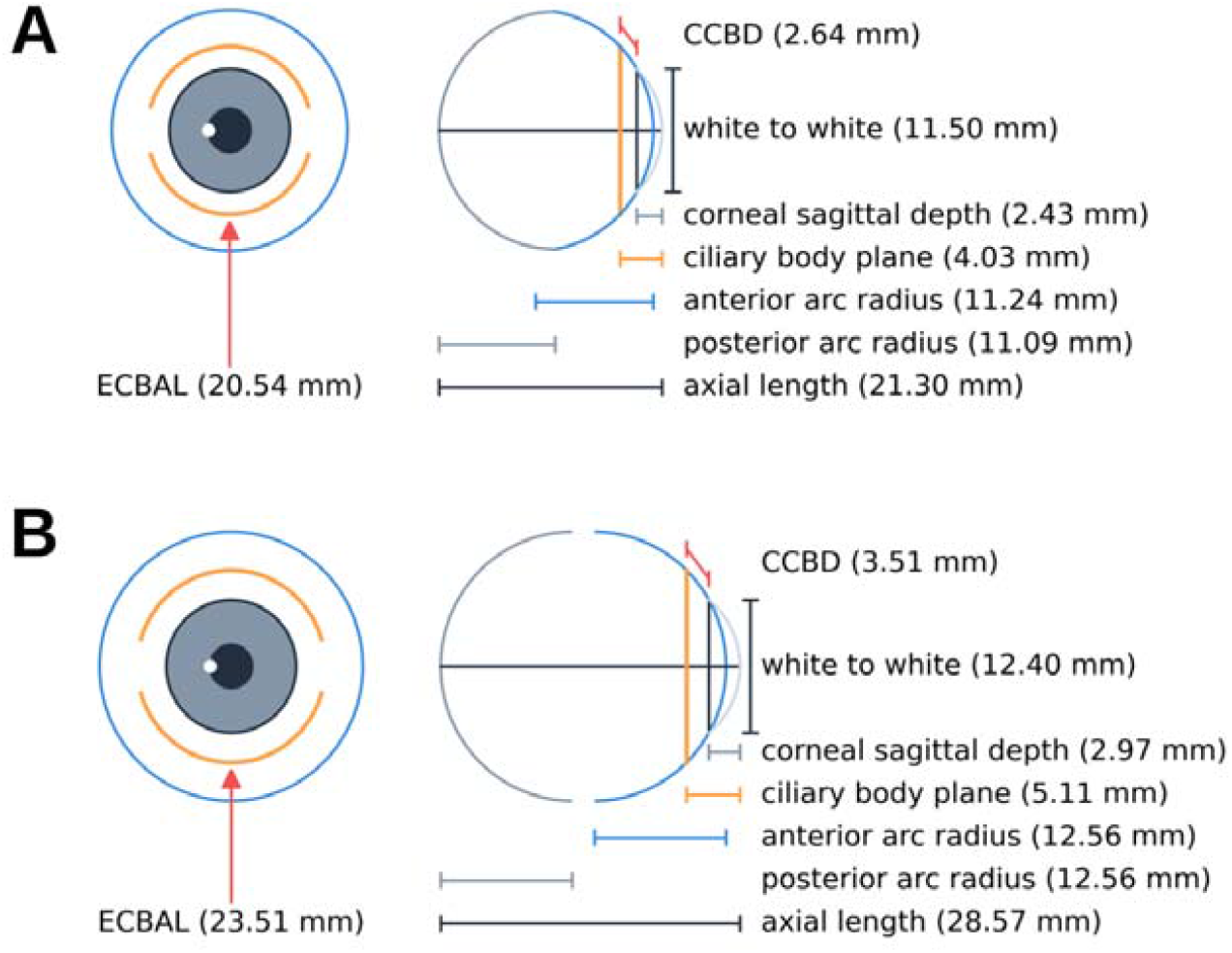
Representative plots of a short (A) and of a long eye (B), numeric values in millimetres represent the lengths of each line segment within the modeled eye; ECBAL (estimated ciliary body arc length), CCBD (calculated ciliary body distance).

AL was defined as the sagittal length of the modeled eye, and the horizontal/vertical diameter was calculated based on linear correlations with AL determined by Jonas et al. from 135 enucleated globes.^10^ This diameter defined an arc of a circle forming the posterior pole, similar to the refractive eye models defined by Emsley, Gullstrand, and Le Grand.^11^

The distance of the WTW from the center of the cornea was calculated from the corneal sagittal depth, which was derived from the mean K, with an asphericity of −0.25. The anterior sclera was defined by another arc of a circle fitted to attach to the limbal endpoints defined by WTW. The radius of this circle was maximized at the radius of the posterior pole to maintain the diameter–AL relationship defined by Jonas et al.^10^ In cases where this maximal radius was reached or exceeded, the posterior arc was not utilized for further calculations. For aesthetic purposes, an arc is also fitted to the WTW and the endpoint of AL to estimate the cornea; however, this arc was not utilized for calculations.

The CB plane was positioned at a distance of ACD + 0.39 × LT from the center of the cornea, where 0.39 is the average ratio of the anterior and posterior LT based on anterior optical coherence tomography measurements.^12^ This plane was projected to the anterior arc to determine the inner diameter of the CB. This diameter formed the basis for calculation of the CB external diameter, 5/12 of which was defined as the ECBAL. The chord distance between the intersections of the WTW and CB plane on the anterior arc was considered the CCBD. The Python script used for these calculations (See Supplementary Material S1) is openly available under the GPLv3 license.^13^

### Model testing

Publicly accessible biometric measurements (AL, K, ACD, LT, and WTW) for 24,001 eyes were pooled to form the study dataset (See Supplementary Material S2). ECBAL and CCBD were then calculated from these data. Descriptive statistics and anonymized data from the study participants are referenced in their respective articles (Table 1).

**Table 1:** Geograhical distribution of the included public datasets.

| Individual eyes in dataset | Eyes included in study | Geographical region |
| --- | --- | --- |
| 150 | 139 | Italy<sup>14</sup> |
| 509 | 505 | USA<sup>15</sup> |
| 2031 | 2031 | USA<sup>16</sup> |
| 266 | 266 | USA<sup>17</sup> |
| 19472 | 19470 | Germany/Austria<sup>18</sup> |
| 175 | 102 | South-Korea<sup>19</sup> |
| 1005 | 1005 | China<sup>20</sup> |
| 118 | 118 | South-Korea<sup>21</sup> |
| 159 | 158 | China<sup>22</sup> |
| 140 | 140 | China<sup>23</sup> |
| 69 | 67 | Thailand<sup>24</sup> |
| 24094 | **24001** | **Total** |

### Fluence calculations

Fluence is calculated as power × duty cycle × dwell time / probe area. Dwell time is dependent on arc length, probe diameter, and sweep time, as detailed in Grippo et al.^4^ For our model, we used a probe diameter of 700 µm and the calculations of dwell time and probe area were rounded to four decimals. Sweep time was set to a constant of 20 seconds, as this pacing seems optimal for continuous manual movement and the 80 second exposure duration can be comfortably split into four sweeps per hemisphere.

### Statistical analysis

Continuous data are reported as the mean ± 2 SD. Eyes with an ECBAL of 21.7– 22.0 mm and a CCBD of 3.75–3.85 mm were further analyzed. Pearson’s correlation was used to assess the relationship between each variable and ECBAL and CCBD. Correlations were considered strong and statistically significant when r > 0.7 and p < 0.05. All analysis was performed using custom Python scripts depending on the scipy, numpy, and matplotlib libraries.

## Results

Table 2 presents the mean biometric data of the pooled dataset. The mean ECBAL was 23.99 ± 1.8 mm. The correlations of ECBAL with AL (Figure 2), K, ACD, LT, and WTW were r = 0.723, –0.367, 0.754, –0.17, and 0.399 respectively (all p < 0.01). The association between ECBAL and AL was y = 0.56x + 10.76 for eyes < 24 mm and y = 0.13x + 21.53 for eyes ≥ 24 mm. The mean CCBD was 4.21 ± 0.8 mm. The correlations of CCBD with AL, K, ACD, LT, and WTW (Figure 3) were r = 0.503, – 0.111, 0.587, –0.078, and –0.162, respectively (all p < 0.01).

**Table 2.** Mean biometric data of the pooled database for all eyes, eyes with 22 mm ECBAL (estimated ciliary body arc length), and eyes with 3.8 CCBD (calculated ciliary body distance) - AL (axial length) ACD (anterior chamber depth), LT (lens thickness), WTW (white to white), and Kmean (mean keratometry in diopters)

|  | All eyes | Eyes with 22 mm ECBAL | Eyes with 3.8 mm CCBD |
| --- | --- | --- | --- |
| AL (mm) | 23.8 ± 3.2 | 21.97 ± 1.7 | 22.95 ± 2.0 |
| ACD (mm) | 3.14 ± 0.9 | 2.36 ± 0.5 | 2.86 ± 0.7 |
| LT (mm) | 4.57 ± 1.0 | 4.59 ± 0.9 | 4.68 ± 0.9 |
| ECBAL (mm) | 23.99 ± 1.8 | 21.87 ± 0.2 | 23.24 ± 1.0 |
| CCBD (mm) | 4.21 ± 0.8 | 3.27 ± 0.5 | 3.8 ± 0.1 |
| WTW (mm) | 11.96 ± 0.8 | 11.62 ± 0.9 | 12.0 ± 0.8 |
| Kmean (D) | 43.87 ± 3.2 | 45.17 ± 3.2 | 44.11 ± 3.1 |

**Figure 2.**
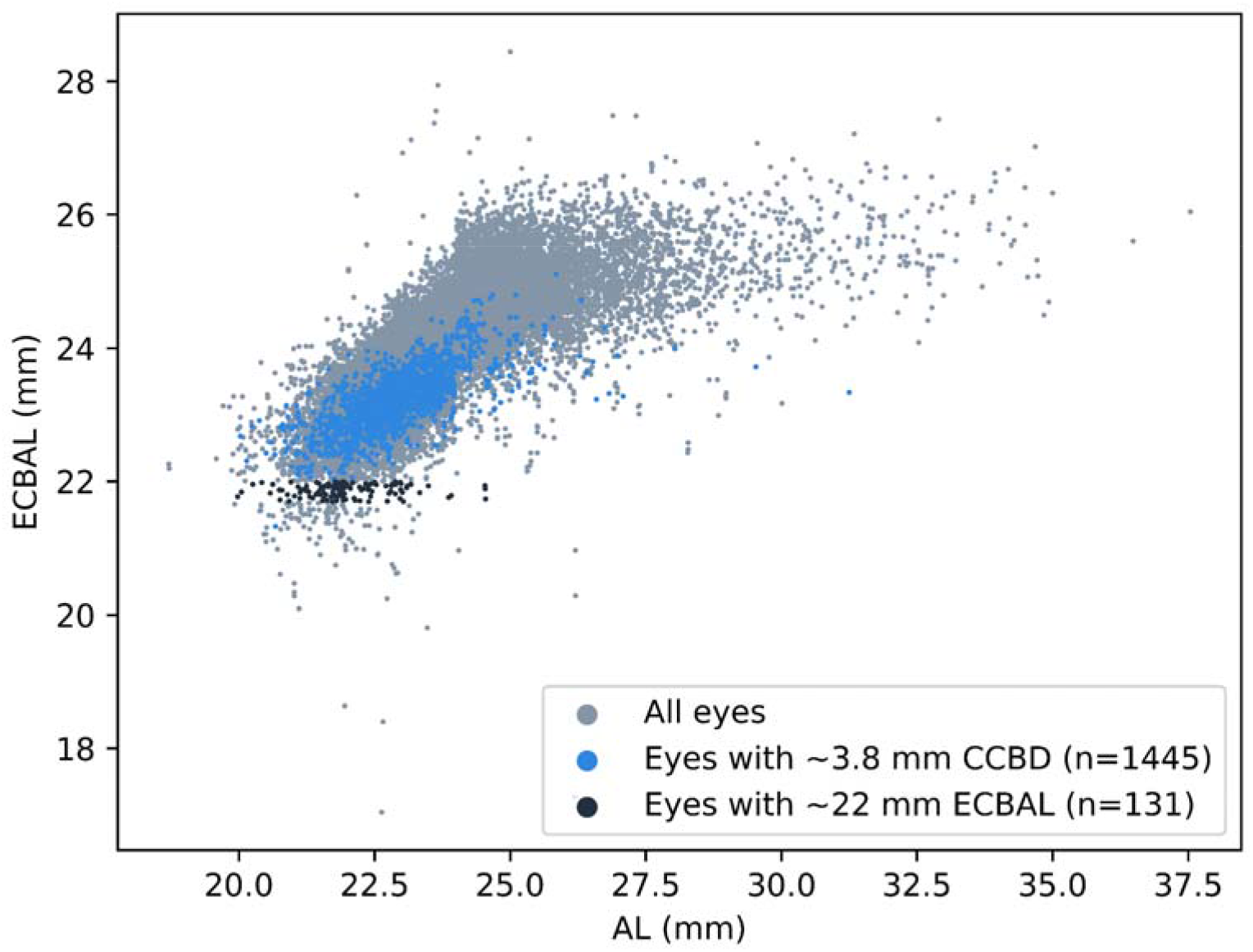
estimated ciliary body arc length (ECBAL) shows strong correlation with axial length (AL). CCBD stands for calculated ciliary body distance

**Figure 3.**
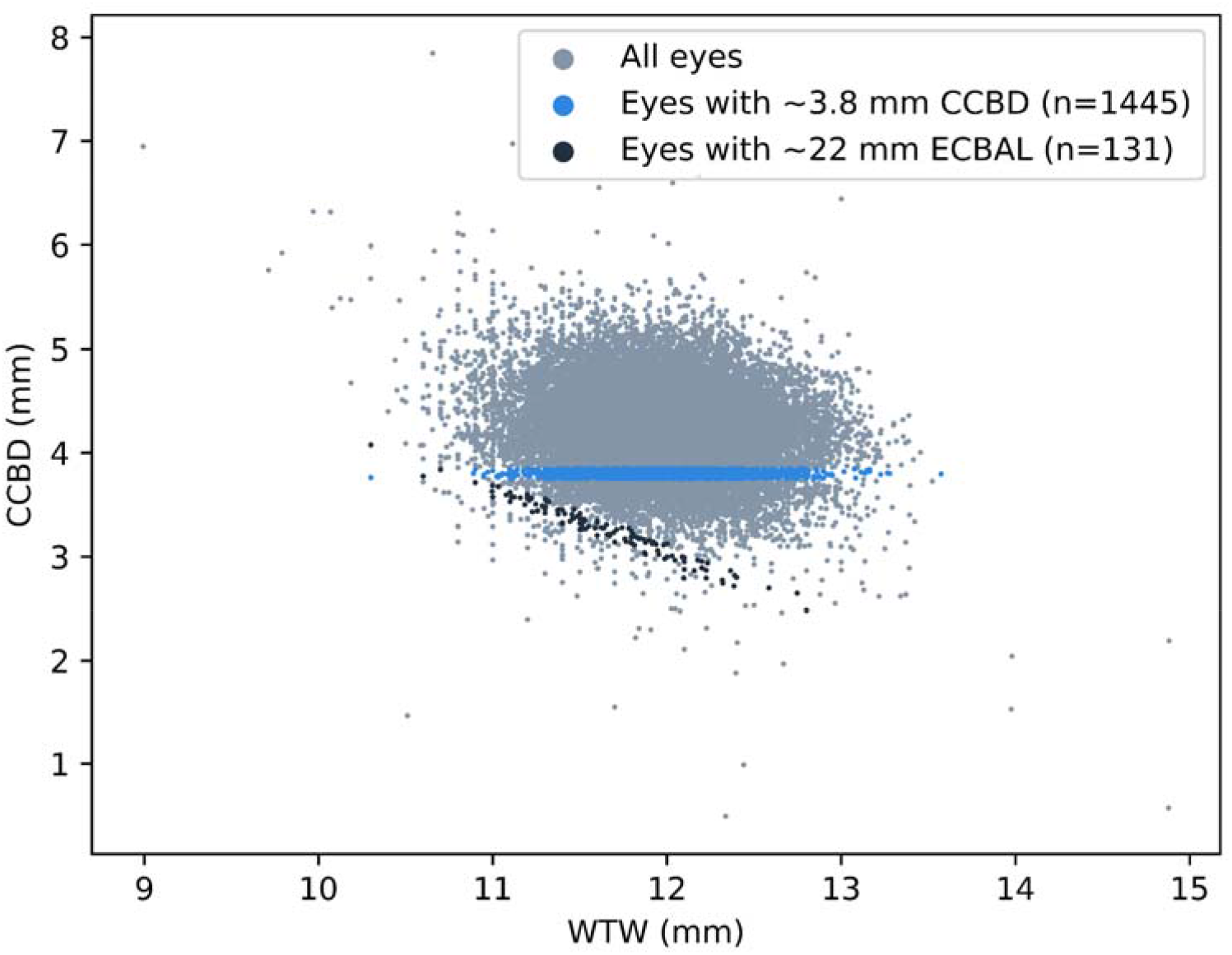
calculated ciliary body distance (CCBD) shows weak correlation with white to white, (WTW). ECBAL stands for estimated ciliary body arc length

AL-based subgroup clustering was performed as follows: very short (<20 mm), short (20–22 mm), average (22–24 mm), and long (>24 mm). ECBAL and CCBD values for each subgroup are shown in Table 3. The number of eyes with an ECBAL 21.7– 22.0 mm was 131 of 24,001 (0.55%); AL and ACD for this subgroup ranged from 19.97–24.55 mm and 1.85–3.36 mm, respectively. The number of eyes with a CCBD of 3.8 mm was 1,445 of 24,001 (6.02%); AL and ACD for this subgroup ranged from 20.02–31.25 mm and 1.86–4.09 mm, respectively. Mean fluence was 120.33 ± 9.0 J/cm^2^, with a maximum and minimum value of 169.14 J/cm^2^ and 101.35 J/cm^2^, respectively. Across the entire dataset, the mean difference of fluence was −8.18 ± 6.9%, ranging from −22.66% to +29.07% in energy density compared to treating only the recommended 22 mm.

**Table 3.** Comparison of estimated ciliary body arc length (ECBAL) and calculated ciliary body distance (CCBD) between eyes of varying axial length.

|  | **Very short**<br>(n=13) | **Short**<br>(n=1,786) | **Average**<br>(n=13,694) | **Long**<br>(n=8,508) |
| --- | --- | --- | --- | --- |
| ECBAL (mm) | 22.44 ± 1.0 | 22.74 ± 1.2 | 23.63 ± 1.2 | 24.85 ± 1.2 |
| CCBD (mm) | 3.71 ± 0.6 | 3.81 ± 0.7 | 4.1 ± 0.7 | 4.48 ± 0.7 |

## Discussion

A mere 0.5% of eyes from the tested dataset had an ECBAL of 21.7–22.0 mm, which is the range in the original fluence article. Moreover, AL and ACD varied widely within this subgroup. Our model confirms that the reference publication^4^ reports measurements from a random eye and that smaller eyes will most likely present with a shorter ECBAL and larger eyes with a longer ECBAL.

We found that ECBAL and CCBD correlated strongly with ACD and AL, rather than WTW. Challenges locating and treating CB is not a new issue.^25^ The ability to predict CB position with calipers set at a specific distance (3.0–3.8 mm) from the limbus is debated, although this is widely done in clinical settings. Notably, the use of transillumination to determine CB dimensions may provide better treatment results.^26^ Our model is not a substitute for the role of transillumination in ST-TSCPC surgery; rather, it shows that CCBD aligns well with histological^27^, ultrasound^28^, and clinical findings^29^, despite contradictory data on the variability of actual CB position and extent.

Reports further suggest that the posterior CB of adult mammalian eyes has substantial secretory activity.^30^ Effects of a more posteriorly placed cyclopotocoagulation^31^ or diathermy^32^ on IOP lowering have been described, and some of the energy may be transmitted to the posterior pole of the eye.^33^ Because the considerable variation in CCBD and ECBAL reflects CB anatomy, fixed energy treatment may result in alterations of fluence up to 50% in different scenarios.

A major limitation of this study is the underrepresentation of eyes <20 mm in the dataset. Generalizability may also be limited, as the majority of subjects were of Caucasian and Chinese ethnicity. There was a limited number of other Asian participants and the proportions of African-American and Latino participants in databases from the USA are unknown.

Because our model corrects for nonsphericity resulting from longitudinal myopic elongation, the steepness of ECBAL growth greatly flattens above AL > 24 mm. This corresponds well with horizontal and vertical globe diameter measurements in anatomical studies.^10^ The model may be more likely to have large error outputs for eyes with atypical biometric values, which has also been observed for intraocular lens power calculation formulas.^34^ Ideally, it would be possible to substitute variables in the Barrett Universal II formula^35^ to obtain the results. Despite its widespread and reliable use in refractive surgery^36^, the exact mathematics supporting prediction of the CB plane or refinement of the estimated lens position are publicly available. Barrett describes the need to build a theoretical refractive model “where the ciliary plane is determined as the intersection of an anterior sphere—related to the radius of the cornea—and a posterior sphere—related to the radius of the globe”.^37^ In our anatomical approximation model, the level of the intersecting spheres is positioned in the WTW plane and the CB is located posteriorly. Notably, due to its rotational symmetry, our proposed model does not account for horizontal elongation of the eye. However, this does not undermine the importance of employing ECBAL as a variable rather than a constant. Future studies are needed to establish connections with existing methods and enhance applicability. It is also imperative to relate calculations to real-life measurements—preferably transillumination—due to the lack of large-scale studies.^29^

Despite published consensus guidelines, SL-TSCPC remains inconsistent in terms of energy^38^ velocity^39^, treatment pattern, and area.^40^ There is growing interest in alternative approaches, such as returning to continuous wave procedures with slow coagulation.^41,42^ Automation of SL-TSCPC laser treatment has been patented (Publication Number WO/2024/127108) by Belkin Vision Ltd. (13 Gan Rave, P.O.Boc 13254, 8122214, Yavne, Israel) and is based on static delivery of energy through a contact lens with protrusions to the target area. If alternative technologies that integrate probe movement and/or target area localization are developed, knowledge of individual CB parameters will become crucial for achieving precise energy delivery.

Overall, this study has demonstrated the need to revise the uniformly applied arc length of 22 mm and incorporate calculations of higher quality or ECBAL assessment to further personalize and standardize SL-TSCPC treatment.

## Conclusions

Although standardized fluence greatly contributes to the success of SL-TSCPC, it is sensitive to various elements, particularly ocular dimensions. This study has demonstrated that use of 22.0 mm as the standard treatment arc length may under- or overdose laser treatment in many eyes. Precise estimation and/or exact localization of the CB treatment area is required to accurately calculate fluence, particularly because human factors—like manual movement and the aim of the probe—are less predictable. Future automation of SL-TSCPC technology may benefit from including this variable.

## Supporting information

Supplementary S1: Python code for ciliary body length and position calculations

Supplementary S2: Dataset of 24,001 eyes

## Data Availability

All data produced are available online, it is pooled from the following openly available sources:
The study pooled parts of the data from the following openly available datasets:
https://doi.org/10.6084/m9.figshare.19932176.v1
https://doi.org/10.6084/m9.figshare.25205159.v1
https://doi.org/10.6084/m9.figshare.26395018.v1
https://doi.org/10.6084/m9.figshare.24852318.v1
https://doi.org/10.6084/m9.figshare.28399544.v1
https://pmc.ncbi.nlm.nih.gov/articles/instance/7775111/bin/pone.0244590.s001.xlsx
https://pmc.ncbi.nlm.nih.gov/articles/instance/10449110/bin/pone.0289033.s001.xlsx
https://pmc.ncbi.nlm.nih.gov/articles/instance/8096100/bin/pone.0251152.s001.xlsx
https://pmc.ncbi.nlm.nih.gov/articles/instance/11253327/bin/12886_2024_3546_MOESM1_ESM.xlsx
https://pmc.ncbi.nlm.nih.gov/articles/instance/6136745/bin/pone.0203677.s001.xlsx
https://pmc.ncbi.nlm.nih.gov/articles/instance/8936461/bin/pone.0265844.s001.xlsx

https://gitlab.com/ThomasHastings/ciliary_body_calc/-/raw/main/dataset.csv

## Acknowledgements

Not applicable.

