## Supplementary S1: Python code for ciliary body length and position calculations for "Paving the way for automated transscleral cyclophotocoagulation: predicting ciliary body arc length from biometric data using a two-sphere eye model"

```
import math
import numpy as np

def distance(x1, y1, x2, y2):
    return math.sqrt((x2 - x1)**2 + (y2 - y1)**2)

def fit_circle(points):
    points = np.asarray(points)
    x = points[:, 0]
    y = points[:, 1]

    # Linear system: x^2 + y^2 + Dx + Ey + F = 0
    A = np.column_stack([x, y, np.ones_like(x)])
    b = -(x**2 + y**2)

    D, E, F = np.linalg.lstsq(A, b, rcond=None)[0]

    x0 = -D / 2
    y0 = -E / 2
    r = np.sqrt(x0**2 + y0**2 - F)

    return x0, y0, r

def corneal_sagittal_depth(wtw, kmean, Q=-0.25):
    R = 337.5 / kmean
    r = wtw / 2
    term = 1 - (1 + Q) * (r**2 / R**2)

    if term <= 0:
        raise ValueError("Invalid geometry: sqrt term ≤ 0. Check WTW, Kmean, and
Q.")

    return (r**2) / (R * (1 + math.sqrt(term)))


def get_ecbal(al, acd, lt, wtw, kmean):
    if al <= 24:
        d_postcircle = 12.8 + 0.44 * al
    else:
        d_postcircle = 19.7 + 0.19 * al
    r_postcircle = d_postcircle/2

    # draw limbus line
    s = corneal_sagittal_depth(wtw, kmean)
    wtw_x = al-s
    wtw_y = wtw/2

    # fit front circle
    front_circle_points = np.array([(wtw_x, wtw_y),
                                    (wtw_x, -wtw_y),
                                    (r_postcircle, r_postcircle),
                                    (r_postcircle, -r_postcircle)])
    xc, yc, rc = fit_circle(front_circle_points)


    # check if the anterior circle ends up larger than the posterior circle
    if xc>r_postcircle and rc>r_postcircle:
        # find the circle based on r_postcircle
```

```
        xd = math.sqrt(r_postcircle**2-(wtw/2)**2)
        xc = wtw_x-xd
        yc = 0
        rc = r_postcircle

    t = np.linspace(0, 2*np.pi, 400)
    x = xc + rc * np.cos(t)
    y = yc + rc * np.sin(t)

    # find cb plane diameter in anterior circle
    distance_from_apex = al-acd-lt/2
    distance_from_circlecenter = distance_from_apex-xc
    cb_radius_circle = math.sqrt(rc**2-distance_from_circlecenter**2)
    cb_point_on_circle = (distance_from_apex, cb_radius_circle)
    cb_distance = abs(distance(distance_from_apex, cb_radius_circle, wtw_x, wtw_y))
    treated_from_circle = 5 * 2 * cb_radius_circle * math.pi / 12
    return treated_from_circle, cb_distance

if __name__ == '__main__':
    print(get_ecbal(23,3, 3.5, 11, 43))
```
